# Social inequalities in COVID-19 vaccine acceptance and uptake for children and adolescents in Montreal, Canada: a cross-sectional study

**DOI:** 10.1101/2021.05.08.21256831

**Authors:** Britt McKinnon, Caroline Quach, Ève Dubé, Cat Tuong Nguyen, Kate Zinszer

**Affiliations:** Centre for Public Health Research, University of Montreal, Montreal, Canada; Dalla Lana School of Public Health, University of Toronto, Toronto, Canada; Department of Microbiology, Infectious Diseases, and Immunology, University of Montreal, Montreal, Canada; Centre de recherche du CHU de Québec, Laval University, Quebec, Canada; Direction régionale de la santé publique de Montréal du CIUSSS du Centre-Sud-de-l’Île-de-Montréal, Montreal, Canada; School of Public Health, University of Montreal, Montreal, Canada

**Keywords:** COVID-19, child health, social inequalities, vaccination

## Abstract

**Background:** The success of current and prospective COVID-19 vaccine campaigns for children and adolescents will in part depend on the willingness of parents to accept vaccination. This study examined social determinants of parental COVID-19 vaccine acceptance and uptake for children and adolescents.

**Methods:** We used cross-sectional data from an ongoing COVID-19 cohort study in Montreal, Canada and included all parents of 2 to 18-year-olds who completed an online questionnaire between May 18 and June 26, 2021 (n=809). We calculated child age-adjusted prevalence estimates of vaccine acceptance by parental education, race/ethnicity, birthplace, household income, and neighbourhood, and used multinomial logistic regression to estimate adjusted prevalence differences (aPD) and ratios (aPR). Social determinants of vaccine uptake were estimated for the vaccine-eligible sample of 12 to 18 year-olds (n=306).

**Results:** Intention to vaccinate children against COVID-19 was high, with only 12.4% of parents unlikely to have their child vaccinated. Parents with younger children were less likely to accept vaccination, as were those from lower-income households, racialized groups, and those born outside Canada. The percent of parents whose child was vaccinated or very likely to be vaccinated was 18.4 percentage points lower among those with annual household incomes <$100,000 vs. ≥$150,000 (95% CI: 10.1 to 26.7). Racialized parents reported greater unwillingness to vaccinate compared to White parents (aPD=10.3; 95% CI: 1.5, 19.1). Vaccine-eligible adolescents from the most deprived neighbourhood were half as likely to be vaccinated compared to those from the least deprived neighbourhood (aPR = 0.48; 95% CI: 0.18 to 0.77).

**Interpretation:** This study identified marked social inequalities in COVID-19 vaccine acceptance and uptake for children and adolescents. Efforts are needed to reach disadvantaged and marginalized populations with tailored strategies that promote informed decision making and facilitate access to vaccination.

## INTRODUCTION

Health Canada approved Pfizer-BioNTech’s COVID-19 vaccine for 12 to 15-year-olds on May 5, 2021, and soon afterwards adolescent vaccination campaigns began across the country.^1^ For children under 12, the results of ongoing COVID-19 vaccine trials are not expected until at least fall 2021.^2^ As the pandemic continues to evolve, ensuring high vaccine uptake among adolescents and potentially among children will be critical to reducing transmission, achieving herd immunity, and resuming social and economic functions.^3^ It is likely to have direct benefits—protecting children against long-lasting effects of infection (i.e., long COVID) and rare cases of multisystem inflammatory syndrome and severe pediatric COVID-19—and may also have the indirect benefit of protecting others by reducing transmission.^4^ The success of Canada’s current vaccination campaign for adolescents and of prospective campaigns for younger children will in part depend on the willingness of parents to accept the vaccine for their children in the context where complications of COVID-19 among young people remain rare.^5^

In several countries, including Canada, childhood routine vaccination rates have been shown to differ by socioeconomic status, race/ethnicity, immigration status, geography, and religion. ^6–8^ While little is known yet about the social patterning of COVID-19 vaccine acceptance among parents, a growing body of research demonstrates socioeconomic and racial/ethnic inequalities in COVID-19 vaccine acceptance and uptake among adults in the US, UK and Canada.^9–12^ Inequalities in vaccine uptake have many contributing factors, including inequitable access to vaccination services among disadvantaged populations, language and resource (e.g., computer access) barriers, and greater mistrust of governments and public health agencies among some equity-seeking groups.^11,13^ Given that low-income and racialized communities have been disproportionately impacted by COVID-19 in Canada, it is critical to understand disparities in vaccine acceptance and ensure equitable vaccine uptake for children to avoid exacerbating existing inequities.^14^ The objective of this study was to examine social determinants (education, household income, race/ethnicity, birthplace, and neighbourhood) of COVID-19 vaccine acceptance and uptake for children and adolescents within a cohort of parents in Montreal, Canada.

## METHODS

We used cross-sectional data from the Children and COVID-19 Seroprevalence Study (EnCORE).^15^ EnCORE is a cohort of children and adolescents attending 30 daycares, 22 primary schools, and 11 secondary schools across four neighbourhoods in Montreal, Canada. The neighbourhoods were selected to reflect diversity in terms of geography, cumulative COVID-19 cases, and neighbourhood socioeconomic status.^15^ Parents or guardians (hereafter referred to as parents) of children aged 2-18 attending participating schools or daycares who completed an online questionnaire between May 18 and June 26, 2021 were included. All participants provided informed consent for the survey and ethics approval was received from the research ethics boards of the Université de Montréal and the Centre Hospitalier Universitaire Sainte-Justine.

The questionnaire for parents of participating children collected information on the COVID-19 vaccination status of their child and, for those who reported their child to be unvaccinated, their intention to vaccinate against COVID-19 (very likely, somewhat likely, somewhat unlikely, very unlikely). Parents were also asked to report their reasons for getting their child vaccinated or being likely to vaccinate (e.g., to protect child from getting sick, to protect family members, recommended by government), or their reasons for being unlikely to vaccinate (e.g., don’t trust vaccines in general, don’t think child will get sick from COVID-19, worried about side effects). To examine social determinants of vaccine acceptance, we categorized the outcome into 3 levels: vaccinated or very likely to vaccinate, somewhat likely to vaccinate, unlikely to vaccinate. This was done for two reasons. First, COVID-19 vaccination became available to adolescents (12 years and older) in Montreal at drive-through sites on May 21, 2021, after around 9% of parents had already completed the questionnaire. Second, given the limited sample size, we wanted to conduct multivariable analyses using the entire sample, which is problematic when age perfectly predicts the “vaccinated” level of the outcome. However, as a secondary analysis, we examined vaccine uptake (vaccinated vs. unvaccinated) for the sample of vaccine-eligible adolescents aged 12-18.

Parent’s education was categorized as: less than bachelor’s degree level (including professional/technical qualification, secondary diploma, and no qualification), bachelor’s degree level, and master’s degree or higher level. Annual household income before tax was grouped into 3 categories (<100K, 100-150K, 150K+). Race/ethnicity was based on 2016 Canadian Census question that asked individuals to which group(s) they identify:^16^ South Asian, Chinese, Black, Filipino, Latin American, Arab, White, Southeast Asian, West Asian, Korean, Japanese, and other (specify). Due to small numbers within specific racial/ethnic groups, we generated a binary variable (racialized person vs. white). The racialized person category is equivalent to Statistics Canada’s definition of visible minority as “persons, other than Aboriginal peoples, who are non-Caucasian in race or non-white in colour.”^16^ Place of birth was defined as Canadian-born vs. foreign-born. Finally, neighbourhood accounted for area-level socioeconomic and/or sociocultural differences: West Island and Plateau Mont-Royal are more affluent and educated neighbourhoods; Montreal North is one of the city’s poorest and most racially diverse neighbourhoods; and Mercier-Hochelaga-Maisonneuve is a working-class neighbourhood with nearly one-third of the population living below the poverty line.^17^ Additional covariates included the child’s age and sex (50% of sample was female), the parent’s gender (83% identified as female), and whether someone in the household works in a healthcare field (17%).

The analytic sample for this study included 809 parents. To account for missing data for household income (8.4%) and education (1.4%), we performed multiple imputation by chained equations, which uses an iterative multivariable regression procedure to generate distributions for each variable with missing data that are conditional on all other variables in the imputation models.^18^ A total of 10 imputed data sets were generated.

### Statistical analysis

We summarized age-specific frequencies of child vaccine acceptance (vaccinated, very likely to vaccinate, somewhat likely to vaccinate, unlikely to vaccinate) among parents and examined reasons for vaccine acceptance or refusal. We then calculated age-adjusted prevalence estimates for our 3-level outcome (vaccinated/very likely to get vaccine, somewhat likely to get vaccine, unlikely to get vaccinate) by parental education, income, race/ethnicity, birthplace, and neighbourhood. We used multivariable multinomial logistic regression models to estimate adjusted prevalence differences and prevalence ratios.^19^ For the sample of vaccine eligible adolescents (n=306), we used multivariable binary logistic regression to examine social determinants of vaccination status. Analyses were performed in Stata version 15.0 (StataCorp LP, College Station, TX) and results were pooled across imputed datasets using Stata’s mi estimate procedures.

## RESULTS

At the time of survey completion, parents reported that 13.6% of children had received at least one dose of vaccine, while 59.7% were very likely, 14.3% somewhat likely, and 12.4% unlikely to have their child vaccinated against COVID-19. Figure 1 demonstrates a clear association between parental willingness to vaccinate and age of the child: 3.8% with 15 to 18-year-olds, 12.1% with 9 to 11-year-olds, and 19.1% with 2 to 4-year-olds were unlikely to accept vaccination. The most common reason parents were unlikely to vaccinate was concern over the lack of information about the vaccine’s safety and potential side effects (71%) and the belief their child would not get seriously ill from COVID-19 (36%). Only 3% of parents unwilling to vaccinate their child reported distrust of vaccines in general. Among parents in this sample, 90% were themselves vaccinated with at least one dose, while 7% were likely and only 3% unlikely to get vaccinated (primarily citing concerns over safety and side effects). Less educated, lower income, foreign-born and racialized parents were overrepresented among the parents unlikely to accept vaccination. Twenty-two of the 25 parents unlikely to accept vaccination for themselves were also unlikely to accept vaccination for their child, while the other 3 were somewhat likely to accept.

**Figure 1:**
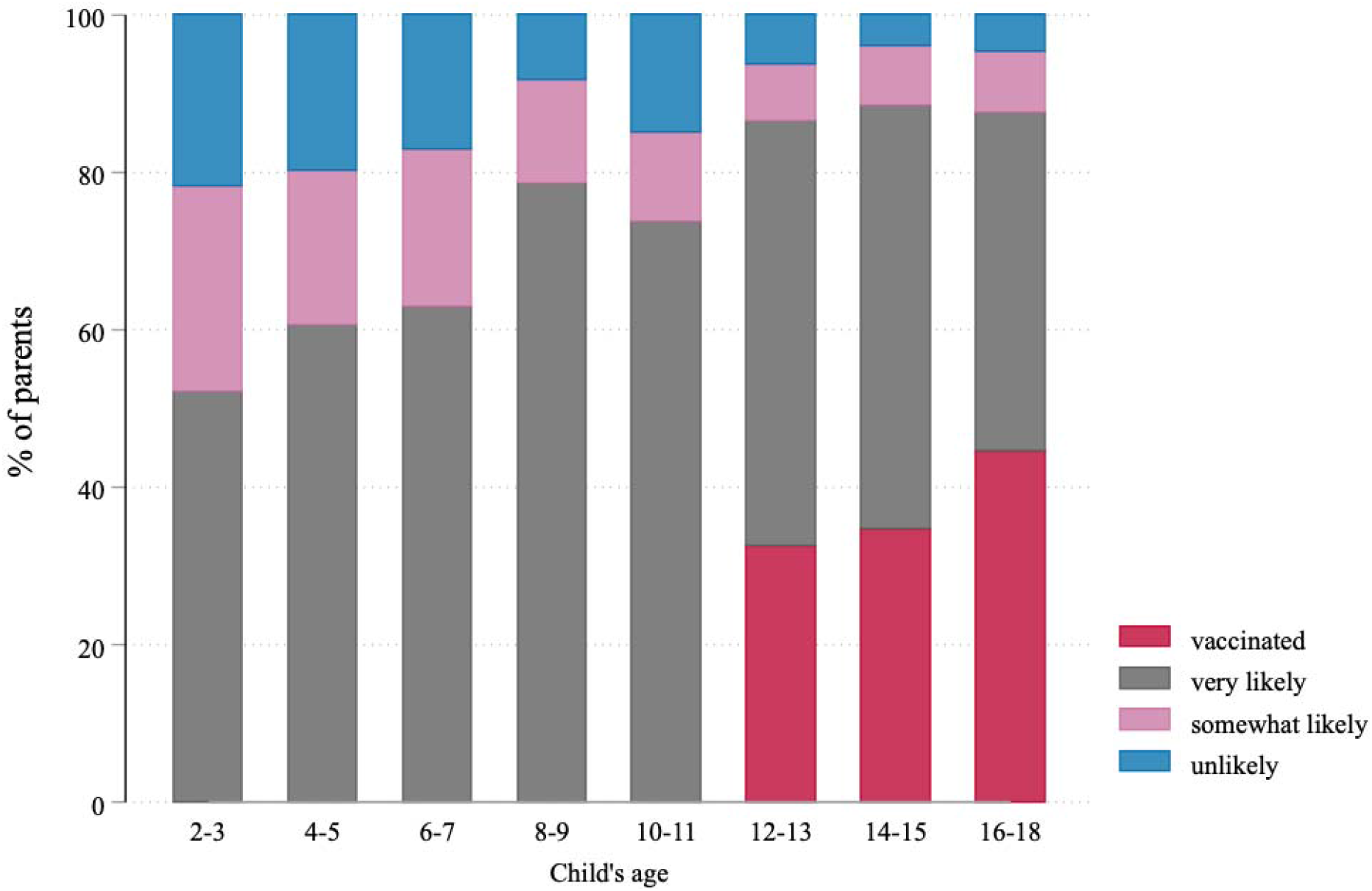
Parent-reported vaccination status and intention to vaccinate by child’s age, May-June 2021

Table 1 presents child age-adjusted prevalence estimates of the parental vaccine uptake/acceptance outcome according to each of the social determinants. Approximately 80% of parents had at least a bachelor’s level of education, 11.6% were classified as a racialized person (comprised of Latin American (35%), Arab (26%), Black (21%), and other (18%)), 23.2% were born outside Canada, two-thirds reported an annual household income >$100,000, and relatively few were from Montreal North (12.4%) compared to the other three neighbourhoods. The prevalence of being vaccinated/very likely to get vaccinated was lower among parents with lower education levels and household incomes, as well as among racialized parents and those born outside Canada. The prevalence of being unlikely to vaccinate was also consistently higher among these groups, as well as among individuals from the Montreal North neighbourhood.

**Table 1:** Prevalence of parent-reported COVID-19 vaccine acceptance for children aged 2-18 according to social determinants, May-June 2021

|  | No. (%)<sup>a</sup> | Child age-adjusted prevalence (95% CI)<sup>b</sup> |  |  |
| --- | --- | --- | --- | --- |
|  |  | Vaccinated or very likely to vaccinate | Somewhat likely to vaccinate | Unlikely to vaccinate |
| Total | 809 | 73 (70, 76) | 14 (12, 17) | 12 (10, 15) |
| Education level |  |  |  |  |
| Less than bachelor's degree | 159 (19.9) | 63 (56, 70) | 17 (10, 23) | 20 (14, 27) |
| Bachelor's degree | 332 (41.6) | 73 (69, 78) | 15 (11, 19) | 12 (8, 15) |
| Master's degree or higher | 307 (38.5) | 79 (75, 83) | 12 (8, 15) | 9 (6, 12) |
| Race/ethnicity |  |  |  |  |
| White | 707 (88.4) | 76 (73, 79) | 14 (11, 16) | 10 (8, 12) |
| Racialized person | 93 (11.6) | 55 (46, 65) | 16 (8, 23) | 29 (20, 38) |
| Place of birth |  |  |  |  |
| In Canada | 620 (76.8) | 78 (75, 82) | 12 (10, 15) | 9 (7, 11) |
| Outside Canada | 187 (23.2) | 57 (51, 64) | 20 (14, 26) | 23 (17, 28) |
| Household income |  |  |  |  |
| <100K | 250 (33.7) | 59 (53, 65) | 22 (17, 27) | 19 (14, 24) |
| 100-150K | 204 (27.5) | 75 (69, 80) | 14 (9, 18) | 12 (8, 16) |
| >150K | 287 (38.7) | 84 (80, 88) | 8 (5, 12) | 8 (5, 10) |
| Neighbourhood |  |  |  |  |
| West Island | 233 (28.8) | 75 (70, 80) | 14 (10, 19) | 11 (7, 15) |
| Mercier-Hochelaga-Maisonneuve | 199 (24.6) | 78 (73, 84) | 12 (7, 16) | 10 (6, 14) |
| Montreal North | 100 (12.4) | 56 (47, 65) | 20 (12, 29) | 24 (15, 32) |
| Plateau Mont-Royal | 277 (34.2) | 74 (69, 79) | 14 (10, 18) | 12 (8, 15) |

Note. CI=confidence interval.
<sup>a</sup> Numbers do not sum to group totals due to responses of "prefer not to answer"
<sup>b</sup> Row percentages may not sum to 100 due to rounding

Table 2 presents fully adjusted prevalence differences (aPD) and adjusted prevalence ratios (aPR) for parental vaccine uptake/acceptance. Disparities according to household income, race/ethnicity, and birthplace remained after adjustment for all social determinants and additional child and parent covariates. The percent of parents whose child was vaccinated/likely to be vaccinated was 18.4 percentage points lower among those with an annual household income <$100,000 vs. >$150,000 (95% CI: 10.1 to 26.7), which corresponds to an aPR of 0.78 (95% CI: 0.68 to 0.87). Parents born outside Canada were less likely to report their child was vaccinated/likely to be vaccinated (aPD=-15.0; 95% CI: - 23.1 to -7.0) and more likely to report being unlikely to vaccinate compared to Canadian-born parents (aPD=7.6; 95% CI: 1.2, 14.0). Racialized parents had twice the prevalence of being unlikely to vaccinate compared to White parents (aPR=2.00; 95% CI: 1.04, 2.95). Lower education level was also consistent with being less likely to vaccinate, although confidence intervals included the null. Neighbourhood differences were not significant after adjustment for the individual level socio-demographic variables.

**Table 2:** Adjusted prevalence differences and ratios of parent-reported COVID-19 vaccine acceptance for children aged 2-18 by social determinants; May-June 2021

|  | Adjusted prevalence differences <sup>a</sup> (95% CI) |  |  | Adjusted prevalence ratios <sup>a</sup> (95% CI) |  |  |
| --- | --- | --- | --- | --- | --- | --- |
|  | Vaccinated or very likely to vaccinate | Somewhat likely to vaccinate | Unlikely to vaccinate | Vaccinated or very likely to vaccinate | Somewhat likely to vaccinate | Unlikely to vaccinate |
| Parent's education level |  |  |  |  |  |  |
| Master's degree or higher | 0 (Ref) | 0 (Ref) | 0 (Ref) | 1 (Ref) | 1 (Ref) | 1 (Ref) |
| Bachelor's degree | -4.2 (-10.5, 2.1) | 2.0 (-3.5, 7.4) | 2.2 (-2.5, 6.9) | 0.95 (0.87, 1.03) | 1.15 (0.71, 1.59) | 1.23 (0.68, 1.79) |
| Less than bachelor's degree | -7.3 (-16.2, 1.7) | -1.0 (-7.8, 6.0) | 8.2 (0.7, 15.6) | 0.91 (0.79, 1.02) | 0.93 (0.43, 1.44) | 1.87 (0.89, 2.85) |
| Parent's race/ethnicity |  |  |  |  |  |  |
| White | 0 (Ref) | 0 (Ref) | 0 (Ref) | 1 (Ref) | 1 (Ref) | 1 (Ref) |
| Racialized person | -6.1 (-15.9, 3.7) | -4.2 (-10.9, 2.6) | 10.3 (1.5, 19.1) | 0.92 (0.79, 1.05) | 0.72 (0.28, 1.16) | 2.00 (1.04, 2.95) |
| Parent's place of birth |  |  |  |  |  |  |
| In Canada | 0 (Ref) | 0 (Ref) | 0 (Ref) | 1 (Ref) | 1 (Ref) | 1 (Ref) |
| Outside Canada | -15.0 (-23.1, -7.0) | 7.4 (0.4, 14.4) | 7.6 (1.2, 14.0) | 0.81 (0.71, 0.91) | 1.60 (0.97, 2.24) | 1.78 (1.01, 2.55) |
| Household income |  |  |  |  |  |  |
| >150K | 0 (Ref) | 0 (Ref) | 0 (Ref) | 1 (Ref) | 1 (Ref) | 1 (Ref) |
| 100-150K | -5.7 (-12.7, 1.2) | 3.9 (-2.0, 9.8) | 1.8 (-3.5, 7.2) | 0.93 (0.85, 1.01) | 1.43 (0.66, 2.21) | 1.20 (0.57, 1.83) |
| <100K | -18.4 (-26.7, -10.1) | 12.7 (5.6, 19.8) | 5.7 (-0.3, 11.7) | 0.78 (0.68, 0.87) | 2.41 (1.19, 3.64) | 1.62 (0.79, 2.45) |
| Neighbourhood |  |  |  |  |  |  |
| West Island | 0 (Ref) | 0 (Ref) | 0 (Ref) | 1 (Ref) | 1 (Ref) | 1 (Ref) |
| Mercier-Hochelaga-Maisonneuve | 6.6 (-1.0, 14.3) | -4.3 (-10.9, 2.2) | -2.3 (-8.2, 3.6) | 1.09 (0.98, 1.20) | 0.72 (0.36, 1.08) | 0.80 (0.36, 1.25) |
| Montreal North | -5.7 (-16.4, 5.0) | 2.3 (-7.1, 11.8) | 3.4 (-4.8, 11.6) | 0.92 (0.78, 1.07) | 1.15 (0.52, 1.78) | 1.29 (0.53, 2.05) |
| Plateau Mont-Royal | 2.3 (-5.2, 9.7) | -2.2 (-8.6, 4.2) | 0.0 (-5.8, 5.8) | 1.03 (0.93, 1.14) | 0.86 (0.48, 1.23) | 1.00 (0.51, 1.49) |

Note. CI=confidence interval.
<sup>a</sup> Prevalence differences, prevalence ratios and 95% CIs were calculated from average marginal effects that were estimated from a multivariable multinomial logistic regression model that included all social factors in the table, as well as child's age and sex, parent's gender, and whether a household member works in healthcare

Approximately one-third (35.9%) of the 306 eligible adolescents had received at least one dose of a COVID-19 vaccine (Table 3). Lower household income and parent’s place of birth outside Canada were consistent with lower vaccine uptake, although 95% CIs slightly crossed the null. Adolescent vaccine uptake in Montreal North was half that for the West Island (aPR=0.48; 95% CI: 0.18, 0.77), after adjustment for parent and household socioeconomic variables.

**Table 3:** Uptake of at least one dose of COVID-19 vaccine among 12-18 year-olds according to social determinants, May-June 2021

|  | % vaccinated with at least one dose | Adjusted prevalence ratio for vaccinated vs. unvaccinated (95% CI)<sup>a</sup> |
| --- | --- | --- |
| Total (N=306) | 35.9 |  |
| Parent's education level |  |  |
| Master's degree or higher | 35.5 | 1 (Ref) |
| Bachelor's degree | 39.2 | 1.15 (0.74, 1.56) |
| Less than bachelor's degree | 31.3 | 1.00 (0.55, 1.43) |
| Parent's race/ethnicity |  |  |
| White | 37.8 | 1 (Ref) |
| Racialized person | 23.7 | 1.06 (0.47, 1.64) |
| Parent's place of birth |  |  |
| In Canada | 39.7 | 1 (Ref) |
| Outside Canada | 23.9 | 0.68 (0.34, 1.02) |
| Household income |  |  |
| >150K | 44.4 | 1 (Ref) |
| 100-150K | 41.2 | 1.07 (0.64, 1.49) |
| <100K | 27.0 | 0.72 (0.41, 1.02) |
| Neighbourhood |  |  |
| West Island | 46.5 | 1 (Ref) |
| Mercier-Hochelaga-Maisonneuve | 34.4 | 0.76 (0.45, 1.08) |
| Montreal North | 19.6 | 0.48 (0.18, 0.77) |
| Plateau Mont-Royal | 35.3 | 0.83 (0.52, 1.14) |

Note. CI=confidence interval.
<sup>a</sup> Calculated from average marginal effects estimated from binary logistic regression models that included child's age and all social factors in the table

## DISCUSSION

COVID-19 vaccine acceptance among the parents of children aged 2-18 was high in this study, with only 12.4% of parents reporting they were unlikely to vaccinate their child. Most hesitant parents reported concerns about the vaccine’s safety and adverse events. Parents with younger children were less likely to accept vaccination, as were those from lower-income households, racialized groups, and those born outside Canada. One-third of vaccine-eligible adolescents had received at least one dose of a COVID-19 vaccine, with those from the most deprived neighbourhood half as likely to be vaccinated compared to the least deprived neighbourhood.

We identified two previous studies that examined parental acceptance of COVID-19 vaccination for children in Canada: one reported 76% of 455 Canadian parents were willing to vaccinate their children^20^ and another from Alberta reported 60% of parents intended, 31% were unsure, and 9% did not intend to vaccinate their child.^21^ This latter study found that families with lower education and income levels reported lower intention to vaccinate their children,^21^ which is consistent with our results showing increased likelihood of vaccination among parents with higher incomes and education levels. These findings for COVID-19 vaccine acceptance for children are also supported by population-based Canadian studies showing that adults with lower education levels,^12,22^ those who self-identify as non-white,^9,12^ and those not born in Canada^22^ had lower intention to receive COVID-19 vaccination. This social patterning also aligns with a substantial body of evidence demonstrating socioeconomic and racial/ethnic disparities in vaccine acceptance and uptake in the United States and the United Kingdom.^11,13^ Just two months into the adolescent vaccine campaign in Canada, little is known about differences in uptake for 12-18 year-olds among equity-seeking groups. However, low-income and ethnically diverse neighbourhoods like Montreal North had the lowest rates of adult COVID-19 vaccination in Quebec as of June 2021. ^23,24^ This parallels our finding that adolescents in Montreal North were around half as likely to be vaccinated as those from the more affluent suburban West Island neighbourhood.

As described above, ample research, including our study, demonstrates the presence of social inequalities in COVID-19 vaccine acceptance and uptake, for both adults and children. Further contextualized research is needed to understand the factors that contribute to COVID-19 vaccine inequalities in children and the types of interventions that can improve COVID-19 vaccination rates and reduce inequities. A systematic review of interventions to reduce inequalities in childhood vaccine uptake found that “locally designed, multicomponent interventions have evidence of effectiveness in urban, ethnically diverse, deprived populations”.^25^ This is in line with the Royal Society of Canada Task Force on COVID-19’s recent recommendation that interventions to improve vaccine acceptance and reduce uptake inequities should be tailored to meet local needs through active engagement and co-development with communities.^26^

Some potential limitations should be considered in interpreting the results of this study. First, we relied on a relatively small convenience sample of parents from four neighbourhoods in Montreal. Compared to 2016 census data, parents comprising the sample had higher education levels, higher median household incomes, and a lower proportion were visible minorities across the four neighbourhoods.^17^ Moreover, the COVID-19 vaccination rate (at least one dose) for our parent sample (90%) was higher than the rate for the population of Montreal aged 20-49 years (71% on June 17, 2020).^23^ Given these differences, we expect estimates of vaccine acceptance for children in our sample may be higher than would be observed in a population-based sample; the reasons for being unlikely to vaccinate may also differ. However, the fact that our sample was relatively socioeconomically advantaged and less ethnically diverse is likely to have resulted in a slight underestimation of inequalities in vaccine acceptance, suggesting the aPD and aPR estimates we present may be conservative.^27^ The limited sample size also did not permit a more granular examination of differences by specific racial and ethnic groups. Among Canadian adults, willingness to accept COVID-19 vaccination has been shown to differ among racial/ethnic groups, with not all racialized groups showing lower willingness to vaccinate.^9,10^ However, in our sample of racialized parents, 82% self-identified as Black, Arab or Latin American, which were the three groups of adults least likely to report vaccine willingness in a national survey.^9^

Racialized and socioeconomically disadvantaged communities have born a disproportionate burden of the COVID-19 pandemic in Canada^14^ and, as of June 2021, low-income and ethnically diverse neighbourhoods in Quebec had the lowest rates of COVID-19 vaccination.^23,24^ Social inequities in vaccination uptake among children will only compound existing inequities. Efforts are needed to reach disadvantaged and marginalized populations with tailored strategies that promote informed decision making and facilitate access to vaccination.

## Data Availability

The datasets generated during the current study will become publicly available upon completion of the study and will be available from the corresponding author on reasonable request.

## Acknowledgements

We gratefully acknowledge the children and parents for their participation in our study along with the different schools, daycares, school boards, and associations for their support.

